# Are clinicians open to less asymptomatic STI screening for chlamydia and gonorrhoea in gay, bisexual and other men who have sex with men and the possibility of not treating positive diagnoses? A qualitative study from Australia

**DOI:** 10.1101/2025.10.15.25337966

**Authors:** Teralynn Ludwick, Tin D. Vo, Lauren Ware, Ethan T. Cardwell, Benjamin Riley, Eric P.F. Chow, Jacqueline Coombe, Daniel Grace, Jane S. Hocking, Fabian YS Kong

**Affiliations:** Melbourne School of Population and Global Health, University of Melbourne, Australia; Factor-Inwentash Faculty of Social Work, University of Toronto, Canada; ASHM Health□; Melbourne Sexual Health Centre, Alfred Health, Melbourne, Australia; School of Translational Medicine, Faculty of Medicine, Nursing and Health Sciences, Monash University, Melbourne, Australia; Centre for Epidemiology and Biostatistics, Melbourne School of Population and Global Health, University of Melbourne, Parkville, Vic, Australia; Melbourne School of Population and Global Health, University of Melbourne, Parkville, Vic, Australia; Dalla Lana School of Public Health, University of Toronto, Canada; Melbourne School of Population and Global Health, University of Melbourne; Australia

**Keywords:** asymptomatic screening, chlamydia, gonorrhoea, MSM, antimicrobial resistance, Australia, sexual health, qualitative

## Abstract

Evidence from real-world studies suggests that 3-monthly screening for asymptomatic chlamydia/gonorrhoea is not reducing incidence and is driving increased antibiotic use and antimicrobial resistance (AMR). While some countries are recommending less screening, changes to guidelines require clinician buy-in. This study explored the views of Australian sexual health clinicians to changing practices around asymptomatic screening for chlamydia/gonorrhoea in gay, bisexual and other men-who-have-sex-with-men (GBMSM) and attitudes to not automatically treating positive diagnoses.

We conducted thematic analysis of 16 semi-structured interviews with clinicians working in a range of settings. Interviews covered: evidence on ‘test and treat’, perspectives on reducing screening and treatment for chlamydia/gonorrhoea in GBMSM, AMR, and managing patient conversations. Clinicians had variable knowledge about the limited effectiveness of asymptomatic screening. Many were open to reduced screening, if provided supporting evidence. Given challenges in reducing medical interventions, they recommended public education to drive changes. While some clinicians supported patient dialogue in treatment decision-making, most felt uncomfortable *not* treating. Concerns included: ideas about their role as doctors; onward transmission (particularly to women); and, complications/uncomfortable symptoms/patient psychological well-being. AMR considerations were less salient.

While the ‘test and treat’ paradigm is engrained, clinicians were open to reduced screening, if provided with clear evidence, but were generally reluctant to not treat. A flexible approach that supports patient empowerment in decision-making about screening frequency and choices around treatment could present a way forward. Changing practice requires education to shift patient/clinician mindsets around what it means to have a positive chlamydia/gonorrhoea diagnosis.

## Background

Rates of chlamydia and gonorrhoea are high among gay, bisexual, and other men who have sex with men (GBMSM), and increasing in many countries, including Australia.^1,2^ Over the last few decades, ‘test and treat’ has been a mainstay of sexually transmitted infection (STI) control. In many high-income countries, guidelines recommend GBMSM screen 3-monthly, including for those on HIV pre-exposure prophylaxis (PrEP).^3-5^ However, some are now questioning the relative utility of frequent screening against potential harms.^6-8^

While there is strong evidence that 3-monthly screening for HIV and syphilis reduces transmission and morbidity,^9,10^ frequent screening for *Chlamydia trachomatis* (CT) and *Neisseria gonorrhoeae* appears to have little impact on the prevalence of these STIs.^11,12^ The recent Gonoscreen study of men-who-have-sex-with-men on HIV PrEP found no evidence that foregoing screening for gonorrhoea and chlamydia in this population is inferior to 3-monthly screening, with no difference in the rate of gonorrhoea infection.^12^ The lack of impact of frequent screening on prevalence in the GBMSM community is largely attributed to high sexual activity within dense sexual networks, leading to a high and sustained equilibrium.^13^

The increased detection of asymptomatic infections resulting from 3-monthly screening has contributed to an associated rise in antibiotic prescribing.^6^ Studies have shown that treating asymptomatic gonorrhoea infections – which pose little harm in cis-males and often naturally resolve within weeks^14,15-17^ – contributes to growing levels of drug-resistant gonorrhoea, now upwards of 50% in some contexts.^18-20^ In addition to threatening the effectiveness of the only remaining, first-line antibiotic for gonorrhoea, and the ability to treat *symptomatic* cases, antimicrobial resistance (AMR) in one organism contributes to resistance in other pathogens.^21,22^

Across the international sexual health community, there remains uncertainty and mixed views on whether to reduce asymptomatic screening.^23,24^ Our study from the Australian context aims to understand the perspectives of sexual health clinicians, including what considerations and concerns underpin their acceptance or resistance to reduced asymptomatic screening and treatment.

## Methods

We recruited Australian clinicians working in sexual health for interviews by: 1. emailing key contacts at major, public sexual health services; 2. embedding an expression of interest within concurrently run surveys on the topic of doxycycline post-exposure prophylaxis (doxy-PEP) and asymptomatic screening that were widely disseminated via peak body networks covering general practice, sexual health and infectious disease. From the large response (>50), we purposively sampled for diversity across geographic location (at least one individual from each of Australia’s states/territories excluding Tasmania for which there were no participants), roles (nurse practitioners (NPs), sexual health physicians, general practitioners (GPs), and public and private practice settings (sexual health clinics, hospitals, Aboriginal health services, LGBTIQ+ organisations).

Semi-structured interviews were conducted online between September-December 2024, with plain language statements and consent forms provided beforehand. Interviews inquired about: knowledge of the evidence on asymptomatic screening, considerations and concerns on reducing screening, and resources needed to support clinicians (Appendix 1). Each interview included two members of the research team (TL, LW, TC, BR) and lasted 30-45 minutes. Participants received a AU$50 gift voucher.

Field notes were taken and initial themes were discussed post-interview. Interviews were digitally recorded and transcribed using an automated service (Otter.ai), and checked for accuracy by LW. Codes and themes were developed inductively by TL in NVivo using reflexive thematic analysis.^25^ The University of Melbourne Human Research Ethics Committee granted ethical approval (2024-28374-58750-7).

## Results

Sixteen clinicians (4 physicians, 8 GPs, 4 NPs) across seven states/territories participated in interviews. Twelve worked in public settings (sexual health centres, hospitals, Aboriginal health services), two in LGBTIQ+ focused non-government organisations, and five in general practice. Most participants had extensive experience with GBMSM clients.

Clinicians acknowledged that ‘test and treat’ has been a mainstay of their practice, but prior to the interviews, most had reflected little on the practice of asymptomatic screening and treatment for chlamydia and gonorrhoea, the evidence behind it, and whether changes were needed. The interviews served as a ‘think aloud’ process.

## Section 1: Perspectives on reducing asymptomatic screening

### 1.1. Willingness to be guided by the evidence

Participants had variable knowledge of the empirical evidence related to frequent asymptomatic screening for chlamydia and gonorrhoea on population prevalence. Some were highly conversant with recent studies, acknowledging that the current 3-monthly recommendation is “*somewhat arbitrarily picked*.’ As such, they were open to changing recommendations while concurrently developing the evidence around the new screening intervals. Others had little familiarity with empirical studies on screening and prevalence, and relied on “seeing the benefit in their individual patient population,” with the assumption that current guidelines were underpinned by supporting evidence. When asked about their knowledge of the evidence on 3-monthly screening, one clinician said:

> *…not having looked at the data myself recently, I’m not entirely sure*…*My hopefully, somewhat, educated guess would be that a ‘test and treat’ policy and having guidelines around how frequently to test certain populations is having an effect on reduced rates*. (GP_7)

Demonstrating that decreased screening intervals would not lead to increased transmission, emerged as a central determinant in shaping the clinician’s level of comfort with reduced screening. Applying a public health lens, clinicians’ primary concern was preventing harm to others (beyond GBMSM), particularly to sexual partners who are women. Consequently, they were very interested in having a research-driven approach, showing the (non)impact of different screening intervals on both asymptomatic and symptomatic infection rates:

> *If I was aware of more concrete evidence that asymptomatic carriage along the community wasn’t causing harm, then I would be open to the idea* [of reduced screening]. *If you could make a comparison, showing that the longer screening durations aren’t resulting in more infections in the community, than evidence such as that would be very useful, because it would make me feel much more comfortable. (GP_8)*

While a minority of participants were more strongly in favour of or against reduced testing, most were uncertain, but expressed a cautious openness to reducing screening frequency. They framed screening frequency as an issue that should be guided by the emerging evidence, and as such were open to being ‘guided by the experts.’

### 1.2 Acceptability to the GBMSM community

Clinicians were concerned about acceptability of reduced screening to the GBMSM community and the potential to cause distress. Clinicians perceived that screening has become a “customary” and “routine” way of the GBMSM “community taking care of each other” and providing “peace of mind”:

> *It’s been so hard drilled until now, that if I told them* [GBMSM clients] *that we’re going to reduce screening frequency…and there’s no clear evidence behind it*… *I’ve just got a few* [clients] *that I’m thinking of who would be terrified. (GP_8)*

Clinicians perceived that reducing interventions for GBMSM communities may be especially hard, given that these communities have been very strong at “self-advocating for access” to sexual health services and reducing access could be perceived as ostracizing:

> *Trying to reduce interventions has always been a complex task*… *Having to go to someone and say, ‘We don’t need to do anything’, that’s really hard*…*because the patient wants it. We don’t want them to feel ostracised or disconnected from sexual health services. (NP_1)*

However, others felt such conversations could be managed through a rational doctor-patient discussion:

> - *Most people are sensible… we have to give people credit; they do manage their own lives. You know, we’re not their mum and dad. Yeah, I think it’s how you sell it to the client. Look, this is the recommendation for screening. We’ll see you every six months. Obviously, if something doesn’t feel right, then this is how you make contact. (NP_11)*

### 1.3 Practical considerations

Resource and time savings were not raised as significant considerations, nor was lost opportunity for patient engagement. With patients already attending clinic on a 3-monthly basis for HIV PrEP, issuing pathology scripts for screening was perceived as fast and easy, without adding to consultation time. Only two respondents raised wider health system costs:

> *I don’t know the actual cost of gonorrhoea and chlamydia screening. So, it would be interesting to see how much it’s actually costing the healthcare system and government…But I suppose, certainly, from a doctor’s time point of view, if they’re already coming in for the script, I kind of figured it’s worth it. (GP_12)*

Clinicians were concerned about the feasibility of delivering differentiated messages on how often to screen for certain STIs over others and for different populations. They noted that this message would be “incredibly hard to manage” and were uncertain “how successful it would be” coming in the face of decades and significant investments in promoting screening. They also worried that providing “wildly different screening suggests for different communities” might make individuals feel like they’re getting “stigmatised by their lifestyle” and risks “pigeon holing people” into different categories.

There was little discussion about the implications of frequent screening on levels of antibiotic consumption.

## Section 2: Perspectives on non-treatment of asymptomatic infections

While many entertained the idea that the sexual health community may be “making a mountain out of a molehill” in terms of the significance of these infections in GBMSM, ultimately, almost every participant landed on the side of treating as the general principle. In considering the acceptability of non-treatment, clinicians raised consideration along three important dimensions, as presented below.

### 2.1 Balancing the clinician’s (felt) obligation to treat versus allowing for patient choice

The question of whether to treat asymptomatic cases sparked clinicians to reflect on their role as doctors and “the way we think and do business” (NP_4). Treating was described as an automatic reflex: “It’s almost knee jerk. Like, oh, positive result, let’s treat” (Physician_5). They recognised how their training has influenced their disposition to treat: “So probably just by teaching, or by the fact that I’m a GP, I will just treat it, because it feels as if you can do something” (GP_10). Many felt “morally obliged” to treat a diagnosed infection:

> *That’s one of the most difficult questions for me in terms of, do you ignore a positive infection? I think once you know it’s positive, you are morally bound to at least offer the treatment (Nurse_11)*

In contrast, some reflected on the “paternalistic approach” and believed that treatment decisions should be determined through shared decision-making with the patient:

> - *“I don’t think I’ve ever had a discussion about, ‘do you want to be treated?’ Very paternalistic*…*I’m so used to just treating any positive result. So, it might be a bit of a shock to the patients…but ethically, it feels a better thing to do, to give the patient an option and to have more autonomy*.*” (Physician_5)*

Some clinicians made parallels to conversations they already have with patients, including those with *Mycoplasma genitalium* (for which antibiotics are unlikely to work) and for sexual contacts where presumptive treatment is no longer the default. For these situations, clinicians noted that some patients opt for treatment and some patients prefer a ‘wait-and-see’ approach:

> - *“I have that conversation with patients all the time…Some people just want to be treated to get it over and done with, whereas the other half are like, ‘Oh, if I don’t need medication, I’d prefer to wait and see’” (GP_12)*

### 2. Balancing the probability of self-limiting infections against potential harms

Several clinicians indicated they had limited or no knowledge about natural resolution, with one respondent noting that no relevant information is presented in key STI reference documents.

Overall, the potential for natural clearance did not significantly impact clinicians’ assessments; they were more concerned about the period of infectiousness and risk to others:

> *I’m not as fussed on natural clearance, probably more curious about transmission rates for people who are asymptomatic*…*I care a lot less about people who have asymptomatic carriage and whether they’re going to naturally clear. I’m more worried about the risk to others (GP_8)*

Clinicians recognised that sexual networks overlap, the gender of sexual partners may change, patients do not always disclose same-sex activity, and can discount what counts as ‘real’ sex (e.g. anal sex). As such, many believed it was protective to treat everyone.

While some mentioned they practice a ‘wait-and-see’ approach to antibiotic treatment in their broader practice (e.g. child most likely sick from a virus; wound management), many differentiated their approach to STI management from these other conditions based on the need to protect others from transmission.

Clinicians were concerned about the potential implications of delayed treatment. From a practical standpoint, some thought it easier to “nip it in the bud” as they figured eventually someone will become symptomatic. Further, many clinicians had seen severe presentations that could have been mitigated with early treatment:

> *I’ve seen too many bad things happen for people who have had those [gonorrhoea] infections… I understand that this is probably not a very rational, scientific explanation that I’m giving you. But I have seen people with really serious gonorrheoa, like it’s gone into their heart valves. It’s sepsis. (Public Health Physician_3)*

By not treating, clinicians felt they would be “unnecessarily increasing psychological distress in patients” related to a “stigmatizing infection”:

> *Whether I’m getting rid of symptoms or not, the diagnosis is what people want to rid themselves of quickly. That you see that really clearly with mycoplasma, where you can explain how limited, how unlikely it is to cause damage, especially in gay men, and yet it can still be just unbearable to have a positive test…If someone is really distressed or worried or is feeling stigmatised, it can be really difficult to convince people not to take antibiotics (Physician_13)*

### 2.3 Balancing two conflicting public health aims - antimicrobial stewardship and STI control

Clinicians were uncertain how to balance two conflicting public health aims – rationalizing use of broad-spectrum antibiotics versus controlling STIs. Clinicians conveyed their strong support for antibiotic stewardship, and the need to use antibiotics judiciously, so they continue to be effective for life-threatening conditions. With this reflection in mind, they noted that urethritis would not meet these criteria:

> *We’re talking about antibiotics that are reserved for life threatening infections, and we’re using them for urethritis. That doesn’t sit well as good antibiotic stewardship either (Physician_13)*

At the same time, they acknowledged that it is not “as black and white as don’t treat the asymptomatic infections” (NP_1), as antibiotic resistance is driven by many factors.

Clinicians were equally concerned that poor commitment to STI control may lead to broader epidemics with serious consequences. They made parallels to how the syphilis epidemic in Australia has grown from a few cases to being declared a Communicable Disease Incident of National Significance:

> - *I suppose it kind of happened with syphilis, didn’t it?*… *If we went back and did it all properly at the start, then it would have been a different conversation. And so, I suppose, with chlamydia and gonorrhoea, if you’re saying it doesn’t matter too much in the MSM community, both crossing over and then it affecting women and fertility, then it does. And so, is it worthwhile treating it now in MSM? (GP_12)*

Connected to this was a concern that a laissez-faire attitude about not treating STIs for GBMSM may damage public health messaging about STIs for the heterosexual population:

> - *‘My friend who’s gay, said, it doesn’t matter if you get chlamydia. It’s not a big deal. Like, so why should I care?’ And it’s like, because you’re a female and you’re going to become infertile, that’s why. The fallout if we stop caring, does that spill across to people that we do need to care about? I’ll summarise by saying I’ve got concerns with that attitude (NP_9)*

At the same time, some clinicians pondered whether it was even feasible to achieve population control and whether we need to relax our approach:

> *We’ve had big conversations about Mycoplasma genitalium and it’s just proving harder and more expensive to treat. So we’ve kind of as a community, just decided it’s not worth it. That conversation has led to, well, should we be rethinking chlamydia and gonorrhoea? (GP_8)*

Given this contentious topic, some noted that making these societal decisions should not be up to clinicians alone, and called for more public input into determining how to balance different public health goals:

> *I think there’s a really strong role for antibiotic stewardship. We cannot afford to blow them. We need to be sensible, but we need to give some of the ownership of those decisions to community. I don’t think that it’s up to us to just make those decisions… And the community need to really understand all sides of the argument (Physician_13)*

## 3. Managing the way forward

When considering changes to screening frequency, clinicians seemed most comfortable with a conservative, incrementalist approach while monitoring the effect of changes:

> *What’s my recommendation? I’d say probably reduce testing for gonorrhoea, chlamydia. I wouldn’t rule it out entirely, because one, patients would not accept that, neither would clinicians (NP_1)*

A few preferred to retain the current recommendations, preferring to “err on the side of caution”, while some wanted to maintain flexibility to make screening recommendations based on individual patient circumstances.

Overall, clinicians were more comfortable with less screening than with not treating asymptomatic cases. The general view seemed to be “if we’re going to test for them, then we have to treat them”:

> - *If we think that asymptomatic gonorrhea is not a problem, either for the individual or for the broader community, then why would we even bother to test for it?*…*Then we wouldn’t be in the dilemma of, do we need to treat it or not?*… *I’m not going to do a test for something, if the only thing I’m going to do with that piece of information is write it down and it doesn’t change my action. Because that, to me, is just a waste of breath (Physician_3)*

## Discussion

While the merits of reduced screening are much debated in the research community, this study is among the first to investigate perspectives of sexual health clinicians – key stakeholders who impact the success of guideline changes. Clinicians’ key concerns centred on risk of transmission and harm to others, while the potential for natural resolution did not feature as a significant consideration. While there were mixed views, overall, most clinician participants were cautiously open to reduced screening, if backed by the evidence showing it does not affect prevalence. However, they were reluctant to adopt a ‘wait-and-see’ approach to treating detected cases. They grappled with how to manage greater patient autonomy and shared decision-making in choices around (non)treatment. Further, clinicians raised practical concerns around community acceptability and managing differentiated screening messages for different populations and STIs.

Our findings point to several strategies that could help address concerns and fears that underlie opposition to reduced screening, including the need for evidence demonstrating no harm, engaging communities in new messaging, and broader societal engagement on AMR. As echoed in Berners-Lee, having confidence and trust in the rationale behind reduced screening was an important theme.^26^ In our study, empirical research presented as the lynchpin underpinning trust and confidence. Given that higher risk of transmission, particularly to women, was a key concern, there is a need to generate and socialise robust evidence which demonstrates that reduced screening will not result in harm.

In our study, clinician’s openness to reduced screening seemed to align with clinicians’ support for evidence-based approaches – the dominant clinical paradigm.^27^ While clinicians often highlight evidence-based medicine as a driver of change, their assessment of the quality of evidence also matters.^28^ As highlighted in this study, clinicians raised the importance of having large-scale studies to provide reassurance that guidelines recommending reduced screening would not result in increased transmission. In practice, however, clinicians can remain conflicted about how to balance the available evidence on population-based screening programs against their desire to ensure their patients do not develop untreated conditions under their care.^29^ In contrast, the idea of not treating patients (despite posing no harm) conflicted with their conception of their role as doctors. Reflecting on why they felt compelled to treat, clinicians in our study referenced the nature of their training and their sense of moral obligation. Changing practice around asymptomatic screening and treatment will require changing clinician (and patient) mindsets about what it means to have a positive chlamydia or gonorrhoea diagnosis and if medical interventions are needed. Supporting patient empowerment in shared decision-making around the benefits and potential harms of treatment, including the option to ‘wait and see’, may help shift conceptualizations about STIs.

Clinicians raised the need for strong community engagement to support informed patient-provider dialogue, to manage community-level messaging, and for public prioritization of AMR. Given that reduced screening goes in opposition to what has been advocated for years, clinicians felt strongly about building community acceptability. Other studies show that considerable fear and resistance to reduced screening exists among GBMSM communities, which is valued for providing psychological safety and has become embedded within GBMSM identity and social standing.^26,30^ Communities should be engaged in shaping new messages that are clear, non-stigmatizing, and do not detract from the importance of STI control.

Further, there is need for broader societal engagement on AMR and how we rationalise antibiotic use. Clinicians were conflicted on what presented the bigger public health risk – the risk of increased AMR posed by frequent screening and treatment, versus the risk of trivializing STIs and generating complacency that leads to future epidemics. Our study participants were clear that it should not be up to clinicians alone to educate patients on AMR, but that communities need to be educated and involved in determining priorities around antibiotic use. Education and public awareness, including resources that better quantify and convey AMR risk, are needed to raise the salience of AMR in the public’s eye and in clinical decision-making.^31^

### Limitations

While the Australian context may differ from elsewhere, it is likely that the value placed on evidence-based approaches and preventing transmission would be echoed internationally by clinicians. Future research should investigate messaging that can strengthen the salience of AMR in decision-making with regards to STIs, and how to strategically communicate new screening messages.

## Conclusion

While the ‘test and treat’ paradigm is strongly ingrained, clinicians were open to reduced screening for chlamydia and gonorrhoea in GBMSM, if provided with clear evidence, but were generally reluctant to stop treating asymptomatic cases. To support practice changes, clinicians need to be reassured with evidence that reduce screening will not result in greater harm. Of equal importance will be the need to shift mindsets among clinicians and GBMSM around what it means to have a positive diagnosis, to determine if medical interventions are needed, and to create space for patient-involvement in those discussions.

## Data Availability

Requests for data will be considered upon reasonable request to the corresponding author.

